# Serological immunochromatographic approach in diagnosis with SARS-CoV-2 infected COVID-19 patients

**DOI:** 10.1101/2020.03.13.20035428

**Authors:** Yunbao Pan, Xinran Li, Gui Yang, Junli Fan, Yueting Tang, Jin Zhao, Xinghua Long, Shuang Guo, Ziwu Zhao, Yinjuan Liu, Hanning Hu, Han Xue, Yirong Li

**Author notes:** Equal contribution and co-first authors (Yunbao Pan, Xinran Li). Author order was determined on the basis of seniority. **Corresponding Authors:** Han Xue, Department of Laboratory Medicine, Zhongnan Hospital of Wuhan University, Wuhan University, No.169 Donghu Road, Wuchang District, Wuhan, 430071, China; Yirong Li, Department of Laboratory Medicine, Zhongnan Hospital of Wuhan University, Wuhan University, No.169 Donghu Road, Wuchang District, Wuhan, 430071, China;.

## Abstract

An outbreak of new coronavirus SARS-CoV-2 was occurred in Wuhan, China and rapidly spread to other cities and nations. The standard diagnostic approach that widely adopted in the clinic is nuclear acid detection by real-time RT-PCR. However, the false-negative rate of the technique is unneglectable and serological methods are urgently warranted. Here, we presented the colloidal gold-based immunochromatographic (ICG) strip targeting viral IgM or IgG antibody and compared it with real-time RT-PCR. The sensitivity of ICG assay with IgM and IgG combinatorial detection in nuclear acid confirmed cases were 11.1%, 92.9% and 96.8% at the early stage (1-7 days after onset), intermediate stage (8-14 days after onset), and late stage (more than 15 days), respectively. The ICG detection capacity in nuclear acid-negative suspected cases was 43.6%. In addition, the consistencies of whole blood samples with plasma were 100% and 97.1% in IgM and IgG strips, respectively. In conclusion, serological ICG strip assay in detecting SARS-CoV-2 infection is both sensitive and consistent, which is considered as an excellent supplementary approach in clinical application.

## Introduction

In December 2019, an outbreak of new coronavirus (SARS-CoV-2, also known as 2019-nCoV) infected pneumonia (COVID-19) was occurred in Wuhan, China and soon spread to other cities and countries. According to the epidemiological analysis conducted by the Chinese Center for Disease Control and Prevention (CCDC), 80.9% of the cases are mild/moderate pneumonia, and the crude overall motility rate is 2.3% (1). The clinical manifestations of most patients include fever, cough, shortness of breath and myalgia etc, and radiographic evidence demonstrated pneumonia with multiple mottling and ground-glass opacity (2). A fraction of patients in serious condition will proceed to complications including acute respiratory distress syndrome (ARDS) and cytokine storm, which may account for the reasons for COVID-19 caused death (3-5). Despite the relatively low fatality, the transmissibility of COVID-19 is proved to be high. Basic reproduction number (*R*_*0*_) of COVID-19 ranging from 1.4 to 6.47, while most of the modeling studies showed the *R*_*0*_ value greater than 3 (6, 7).

SARS-CoV-2 belongs to lineage B of the beta-coronavirus family, the zoonotic-origin single-strand RNA viruses that are transmitted between animals and people. The four prevalent viruses of seven coronavirus family members, 229E, OC43, NL63, and HKU1, cause only the mild upper respiratory diseases, while the other two highly pathogenic strains, SARS-CoV and MERS-CoV, together with the new identified SARS-CoV-2 are regarded to pose the global threats to public health. The full-genome sequencing data from two groups revealed that the SARS-CoV-2 comprises of six major open reading frames (ORFs) and shares approximately 80% of similarity with SARS-CoV, but 98.65% nucleotide identity to partial RdRp gene and 96.2% identity to RaTG13 of SARS-like bat coronavirus strain, respectively (8, 9). Similar to SARS-CoV, SARS-CoV-2 recognizes the same cell entry receptor, angiotensin-converting enzyme II (ACE2). Real-time reverse-transcript Polymerase Chain Reaction (real-time RT-PCR) is regarded as the “gold-standard” in the diagnosis of SARS-CoV-2 (10). Designed primers targeting *ORF1ab*, envelop protein gene (E gene) or nucleocapsid protein gene (N gene) are proved to be both sensitive and specific toward the new SARS-CoV-2 virus and ruled out most of the other coronaviruses (229E, OC43 and MERS) and influenza viruses (H1N1, H3N2, H5N1, and H7N9 subtype) (11, 12).

Despite the real-time RT-PCR is widely adopted as the standard diagnostic method of SARS-CoV-2 in China and worldwide, the limitation of this technology is obvious. The patients that are positive to RT-PCR tests can be diagnosed as SARS-CoV-2 infection, while negative to the tests cannot rule out the possibility. The increasing number of reports from local authorities addressed the unneglectable false-negative rate in the diagnosis of suspected cases, which poses much threat to the community and adds an extra burden to the epidemic prevention. For example, the chest CT scanning from 5 suspected cases demonstrated ground-glass opacity and/or mixed consolidation, yet the initial real-time RT-PCR test showed to be negative. After a few days, all the 5 cases tested for the second or third examinations had eventually confirmed to be positive to SARS-CoV-2 (13). Complementary to RT-PCR, pneumonia radiography provides strong evidence for the diagnosis. Nevertheless, more specific measurements, such as serologic methods are urgently needed in the supplement to the current diagnosis. Here we present the newly developed serological detection methods targeting the viral antibody, colloidal gold-based immunochromatographic (ICG) strip, that conducted in Zhongnan Hospital of Wuhan University, hoping to provide supplementary diagnostic approaches for the SARS-CoV-2 infected patients.

## Methods

### Colloidal gold-based immunochromatographic (ICG) strip assay

The blood samples were collected, and blood serum, plasma or whole blood were subjected to ICG assay in accordant with the manufacturer’s protocol (Zhuhai Livzon Diagnositic Inc.). In brief, 10 μL of serum or plasma, or 20 μL of whole blood samples were added onto the sample loading area followed by 100 μL (2 drops) of sample dilution solution. After a short time (no longer than 15 minutes) of incubation, viral IgM- or IgG-containing positive samples (figure 1) could show up both the T line (test) and C line (control); the samples with only C line were regarded as negative (figure 1); the strips with no C line showed up should be considered as the invalid test.

**Figure 1.**
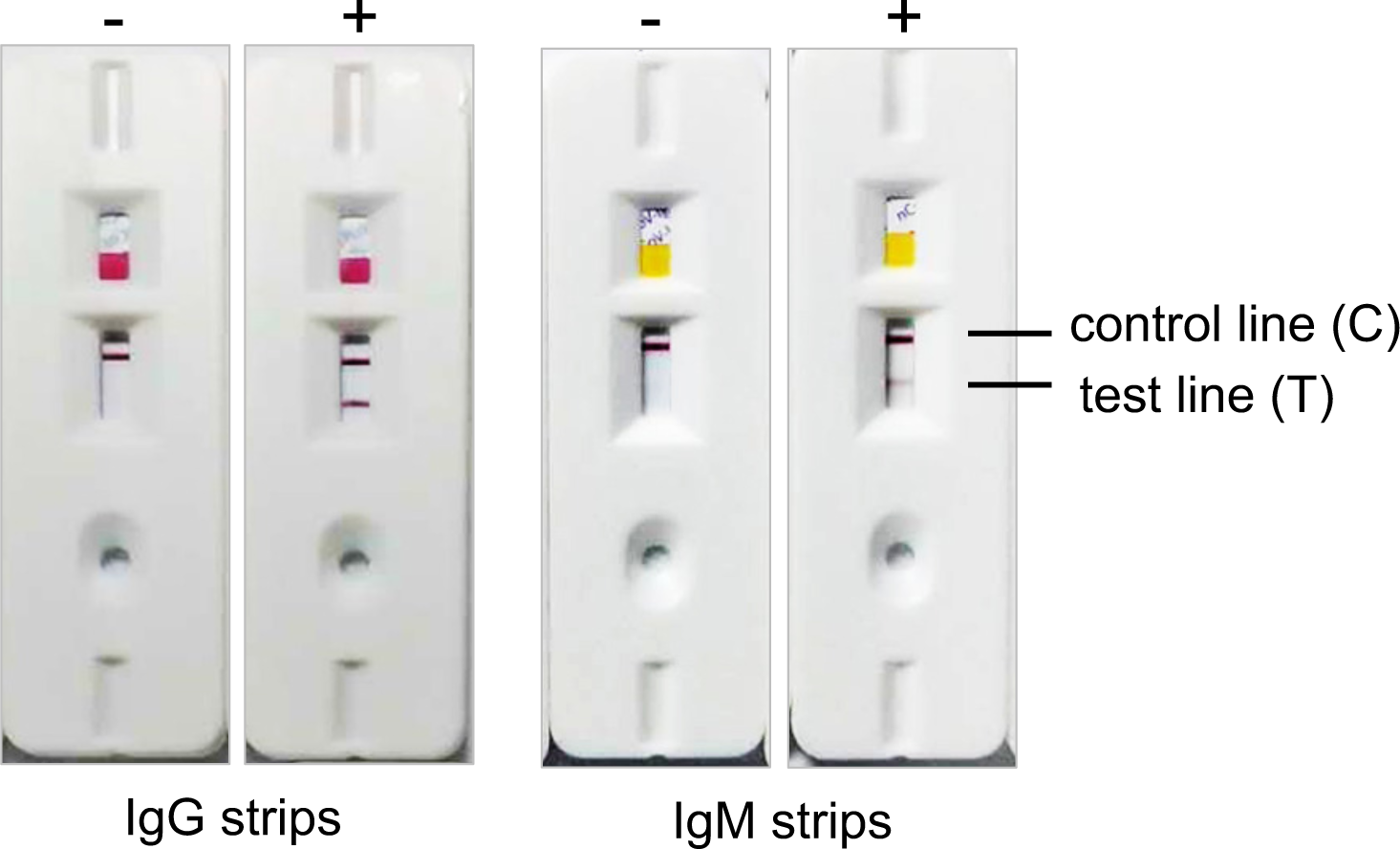
Typical results of the ICG test strip for IgG (A) and IgM (B) antibodies against SARS-CoV-2. The representative results of negative (left) or positive (right) are shown. Upper line in each strip indicates control line (c), lower line in each strip indicates test line (T).

### Real-Time RT-PCR Assay

Throat swab samples were collected and tested for SARS-CoV-2 with the Chinese Center for Disease Control and Prevention (CDC) recommended Kit (BioGerm, Shanghai, China), following WHO guidelines for qRT-PCR. All samples were processed simultaneously at the Department of Laboratory Medicine of Zhongnan Hospital of Wuhan University. All patients with COVID-19 pneumonia tested positive for SARS-CoV-2 by use of quantitative RT-PCR on samples from the respiratory tract. This study was reviewed and approved by the Ethical Committee of Zhongnan Hospital of Wuhan University. Written informed consent was waived by the Ethics Commission for emerging infectious diseases.

### Statistical analysis

In the ICG strip sensitivity assay with real-time RT-PCR confirmed cases and ICG detection capability with nuclear acid negative cases, the numbers of IgM positive, IgG positive and either IgM or IgG positive were counted, and the percentages were presented. In the ICG detection consistency assay, the consistency rate was calculated by the formula: TPR = TP / (TP+FN); TNR = TN / (TN+FN); Consistency = (TP+TN) / (TP+FP+TN+FN). TPR: true positive rate; TP: true positive; TN: true negative; FP: false positive; FN: false negative.

### Data availability

The data that support the findings of this study are available from the corresponding author upon reasonable request.

## Results

### Information of COVID-19 confirmed or suspected patients

A total of 134 samples from 105 patients (48 male vs. 57 female), with a median age of 58 years (range from 20 to 96 years old) that hospitalized at Zhongnan hospital were enrolled in the study. Seventy-eight patients were collected the blood once, 25 patients were collected twice and 2 of the patients were collected three times. Among the samples, 95 of the samples from 76 patients were initially confirmed as SARS-CoV-2 infection by real-time RT-PCR, 8 of the patients were initially negative to nuclear acid detection but positive to the following tests; in total 39 nuclear acid negative samples from 37 of the patients were “clinically diagnosed” as SARS-CoV-2 infection according to the 5^th^ edition of guideline on diagnosis and treatment of the novel coronavirus pneumonia. Specifically, the “clinical diagnosis” means the suspected cases were negative to the real-time RT-PCR test but presented viral pneumonia by radiography. The study was conducted with the consent from all patients and approved by the ethics committee of Zhongnan hospital.

All the blood samples were collected between February 6 and February 21, and the ICG strip assay was performed between February 10 and 23. In total of 108 samples, including 86 of confirmed samples and 22 clinical diagnosed samples, were available for the symptom onset information (disease duration 0-34 days), which was range from January 7, 2020 to February 18, 2020, and subjected to the IgM or IgG sensitivity assay; the symptom onset information was not available for remaining 26 cases. Among all the samples, 39 “clinically diagnosed” samples from 37 patients were subjected to antibody detection capability in nuclear acid negative patients; 47 of the samples were used for the comparison of the consistency in plasma and whole blood samples, 13 of which were ruled out as the samples were expired.

### Sensitivity of IgM or IgG ICG strip assay

A total of 86 samples from 67 cases of real-time RT-PCR confirmed SARS-CoV-2 positive patients with disease duration information were subjected to the analysis. According to the disease progress stage calculated from the start of symptom onset, the disease was divided into early stage (1-7 days from onset), intermediate stage (8-14 days) and late stage (more than 15 days). As summarized in table 1, the positive rates of IgM or IgG in the early stage are relatively low, and gradually increase during the disease progression. The IgM positive rate rising from 11.1% of early stage to 78.6% and 74.2% in intermediate and late stage, respectively. The IgG positive rate in the confirmed patients is 3.6% in early, 57.1% in intermediate and 96.8% in late stage, respectively. Noteworthy, combining the result of IgM and IgG, i.e. patients with either IgM or IgG positive, would significantly increase the sensitivity of IGC assay, especially at the intermediate stage. While IgM and IgG positive rates at the intermediate stage are 78.6% and 57.1%, respectively, combining both parameters would bring a positive rate to 92.9%. Collectively, in response to the virus invade, the IgM was firstly produced against the virus; with the disease progression, IgG antibody start to produce and gradually become detectable. The sensitivity of ICG assays increase and peak at around 15 days. Combining both IgM and IgG would largely sensitize the result of ICG assay.

**Table 1.** Sensitivity of IgM or IgG ICG strip assay in confirmed patients

| Disease duration | No. of sample | No. of IgM+ (%) | No. of IgG+ (%) | No. of IgM+ or IgG + (%) |
| --- | --- | --- | --- | --- |
| 1-7 days | 27 | 3 (11.1%) | 1 (3.6%) | 3(11.1%) |
| 8-14 days | 28 | 22 (78.6%) | 16 (57.1%) | 26 (92.9%) |
| ≥15 days | 31 | 23 (74.2%) | 30 (96.8%) | 30 (96.8%) |
| In total | 86 | 48 (55.8%) | 47 (54.7%) | 59 (68.6%) |

### Antibody detection in nuclear acid-negative “clinically diagnosed” patients

According to the 5^th^ edition of the guideline on diagnosis and treatment of the novel coronavirus pneumonia, highly suspected COVID-19 patients with real-time RT-PCR negative but present radiographic viral pneumonia were regarded as “clinically diagnosed” patients. Although the nuclear acid test is regarded as the “gold standard” of the diagnosis, due to certain limitations, the false-negative cases are not rare. Hence, the IgM and IgG anti-viral antibody were also examined in clinically diagnosed patients. In total 39 samples from 37 clinically diagnosed patients were included. Among these samples, 9 (23.1%) of them were positive to IgM and 15 (38.5%) of them were positive to IgG; when combine the IgG and IgM results in total 17 (43.6%) of samples were positive from 39 nuclear acid negative cases.

Considering the disease stage affects the detection of viral antibody, we also analyzed the clinically diagnosed patients in different stages. As shown in table 2, 22 samples from clinical diagnosis patients with disease duration information were included. 9 cases were at early stage, 6 cases at the intermediate stage and 7 cases at late stage. The positive percentages of IgM at early, intermediate and late stages were 22.2%, 33.3% and 57.1%, respectively; the positive rate of IgG at early, intermediate and late stages were 44.4%, 66.7% and 71.4%, respectively. When it came to IgM and IgG combination, the positive rate boosted to 83.3% in cases at intermediate stage. Comparing with confirmed cases, the IgG and IgM positive rates were similar, albeit relatively lower. In accordant with nuclear acid positive cases, the IgG and IgM combinatorial detection strategy in clinically-diagnosed intermediate-stage patients demonstrated the maximal detection efficacy. In summary, IgM or IgG ICG strip detection in nuclear acid negative suspected patients showed promising detection capability, which would be an excellent supplementary approach in the clinically diagnostic application.

**Table 2.**
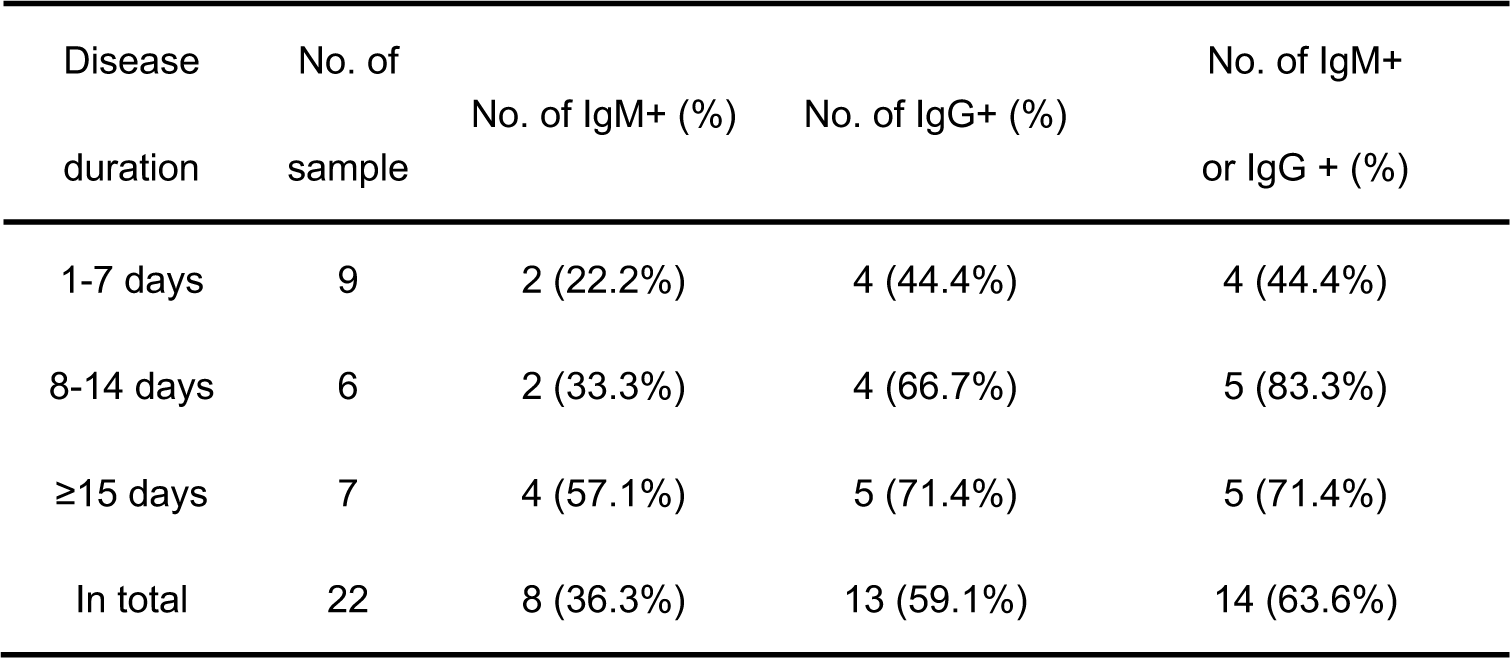
Detection of viral IgM or IgG in real-time RT-PCR negative patients

| Disease duration | No. of sample | No. of IgM+ (%) | No. of IgG+ (%) | No. of IgM+ or IgG + (%) |
| --- | --- | --- | --- | --- |
| 1-7 days | 9 | 2 (22.2%) | 4 (44.4%) | 4 (44.4%) |
| 8-14 days | 6 | 2 (33.3%) | 4 (66.7%) | 5 (83.3%) |
| ≥15 days | 7 | 4 (57.1%) | 5 (71.4%) | 5 (71.4%) |
| In total | 22 | 8 (36.3%) | 13 (59.1%) | 14 (63.6%) |

### ICG assay consistency between whole blood and plasma samples

In order to study the consistency of whole blood samples with plasma, 34 paired whole blood and plasma samples were collected and subjected to IgM or IgG detection. Twenty-four of the plasma positive and 10 of the negative samples were used as a reference. Notably, all the IgM detection in whole blood samples were 100% consistent with plasma samples. In the cases of IgG (shown in table 3), 23 of the plasma positive samples were identified in whole blood samples, which represented the positive consistency rate of 95.8%. As of 10 negative samples in plasma, the whole blood detection of these samples were all negative and demonstrated the 100% negative consistency rate. Compared with plasma antibody detection, the total consistency rate in whole blood ICG assay was 97.1% [(23+10)/(23+0+1+10)]. Collectively, the whole blood antibody detection consistencies were 100% and 97.1% in IgM and IgG ICG assay, respectively, which indicated that the whole blood samples showed excellent agreements with plasma samples.

**Table 3.** IgG detection consistency between whole blood and plasma samples

|  |  | Serum/Plasma |  | In total |
| --- | --- | --- | --- | --- |
|  |  | + | - |  |
| Whole | + | 23 | 0 | 23 |
| Blood | - | 1 | 10 | 11 |
| In total |  | 24 | 10 | 34 |

## Discussion

The late 2019 and early 2020 have witnessed the third and the largest coronavirus outbreak in recent two decades. Due to the limited knowledge related to the new virus SARS-CoV-2 at early stage of the outbreak and the capacity of human-to-human transmissibility in the latent period, the infected cases were exponentially arising in Wuhan, the center of the epidemic, and rapidly spread to domestic and abroad areas. Scientific groups have quickly deciphered the viral whole-genome sequence and specific primers targeting SARS-CoV-2 were designed and tested preclinically before massively diagnostic application in clinic. The viral whole-genome sequencing and viral nuclear acid detection by real-time RT-PCR are regarded as standard diagnostic approaches. While the whole-genome sequence is both time-consuming and labor-intensive, real-time RT-PCR is relatively fast and easy to carry out in hospital laboratories, which makes the latter technique as the “gold standard” of clinical diagnosis. However, the gold standard also has its limitations. Based on more than 3,000 detected cases by real-time RT-PCR in the clinical laboratory of Zhongnan hospital, our previous review summarized several reasons for false-negative incidence, at both detection level and patients level (14). From detection aspects, the quality and sensitivity of detection kits and viral preservation solutions from different companies may all affect the detection accuracy and result in the false-negative possibility. Besides, it is indicated that, during the disease progress, the heavy infection at the nasopharyngeal area of early stage would possibly become negative and the lower respiratory tract may severely be infected at late stage. Thus, the nasopharyngeal swab may not be the best sampling site for all patients at various disease stage. Moreover, patients with persistent anti-viral medication (such as anti-HIV drugs) may decrease the viral loading at the undetectable level. Therefore, additional detection techniques are urgently needed in the supplement to the current diagnostic shortages for the nuclear acid-negative suspected cases, whom would otherwise be ruled out.

In this study, we investigated the quick detection approach targeting viral IgM or IgG antibody, the colloidal gold-based immunochromatographic (ICG) strip assay, in comparison with real-time RT-PCR testing. The antibodies are produced and secreted by B lymphocytes of the adaptive immune system when the foreign pathogens invaded. IgM is usually the first responded antibody that eliminating pathogens before sufficient IgG is produced, while IgG serves as the most robust antibody-based immunity. From the retrospective study of immunoglobulins against SARS-CoV, IgM and IgG were started to be detected after 7 days of onset and persistent for 2-3 years (15, 16). Similar to SARS-CoV, COVID-19 patients also showed similar characteristics. As demonstrated by Zhang et al., both IgM and IgG can be detected after 5 days of onset by anti-SARS-CoV-2 ELISA assay (17). In accordant with *Zhang et al*., our study showed that both IgM and IgG were firstly detected on day 4 among the confirmed cohort and the positive rate of IgM and IgG were 11.1% and 3.6% in early stage patients, respectively. The positive rate of IgM remained approximately 75% in the intermediate and late stage patients, while IgG positive rate kept increasing during the disease progression and up to 96.8% of late-stage confirmed patients showed detectable IgG. Noteworthy, combining IgM and IgG detection results could reach the maximal testing efficacy, especially in intermediate stage. A similar trend could be captured in clinically diagnosed patients. In addition, the consistency rate of whole blood and plasma samples in IgM and IgG assays were 100% and 97.1% respectively, demonstrating an excellent consistency between two types of samples. As the whole blood samples are capable of detecting antibody by ICG strip assay, the quick and massive viral detection in equipment-insufficient laboratories and communities is realizable.

The advantages of ICG strip viral antibody detection are obvious. First of all, the sensitivity of ICG strip is comparable, especially after 7 days of onset; the detection capacity in nuclear acid-negative “clinically diagnostic” patients is impressive. Therefore, ICG strip is highly recommended to be utilized as the supplementary diagnostic approach in clinical applications. The ICG strip can also be widely adopted in the areas where the diagnostic capacity is limited. Secondary, the ICG strip assay is operational as it is ready to use and time-saving. The assay can be finished within 15 minutes without specialized equipment. Thirdly, unlike oral swab sampling that may cause stimulated retching and coughing, which increases the exposure risk of laboratory technicians, the blood collection could avoid the unnecessary risk and reduce the operation steps that may cause aerosol. Fourthly, the detection of antibodies may also indicate the disease recovery, as immunoglobulins are among the most important soldiers in the battle of viruses. Patients that initially detected as virus-positive and gradually become negative but with detectable IgG or IgM during the disease progress may be considered as recovered from this battle. It is noteworthy that ICG strip can only provide qualitative results but the serological ELISA assay against viral antibody offered the quantitative antibody titer and regarded as the superior alternation at this point. Last but not least, ICG strip can be used for community surveillance. Some healthy people in epidemic areas may already get infected but without any symptoms, the detectable antibody from their blood is the evidence of infection. The surveillance detection in epidemic areas for both healthy members and patients could provide useful information for mapping the whole pictures of viral transmission, which would help us to better understand the real situation and establish optimal policies against the epidemic.

Although ICG strip assay is characterized as a rapid and sensitive complementary detection method against SARS-CoV-2, there are several limitations of our test. One is that the assay was carried out without specificity analysis. Our hospital is located in Wuhan, the center of the epidemic, and there are hundreds of, sometimes thousands of, confirmed or suspected COVID-19 cases each day. It is inoperable to select “uninfected” cases for the negative control of specificity analysis, as the patients received after the outbreak may possibly get infected with or without any symptoms. Such assay can be carried out in the areas where the epidemic is not severe and community transmission is at a low chance. Another limitation, as mentioned above, is that the ICG results are qualitative. Despite the positive bands on the strips may provide diverse gradations of color, i.e. deep or light red, the color itself doesn’t correlate with the abundance of antibody. A more precise way of detecting antibody titer is by ELISA assay, though compromised the convenience.

Collectively, we provided a sensitive and consistent serology diagnostic approach in complementary to the current clinically used real-time RT-PCR testing in diagnosis with SARS-CoV-2 infected COVID-19 patients.

## Acknowledgments

This work was supported by the National Key Research and Development Program of China (2018YFE0204500). We acknowledge all health-care workers involved in the diagnosis and treatment of patients in Wuhan.

## Author contribution

Conceptualization: Y. P., Y.L.; Data curation: Y.P., G.Y., J.F., Y.T., J.Z., X.L., S.G., Z.Z., Y.L., H.H.; Formal analysis: Y.P., X.L.; Funding acquisition: Y.L.; Investigation: H.H., Y.L.; Methodology: Y.P.; Supervision: Y.L.; Writing - original draft: X.L.; Writing - review & editing: Y.P.

